# Higher mortality after cardiac surgery is observed in cohort from disadvantaged neighborhoods

**DOI:** 10.1101/2025.07.09.25331232

**Authors:** Mitchell C Haverty, Rittal Mehta, Justus G Reitz, Alyssia Venna, Aybala Tongut, Manan Desai, Monika Goyal, David Wessel, Yves d’Udekem, Jennifer H Klein

## Abstract

**Background:** Socioeconomic disadvantage is linked to adverse outcomes after pediatric cardiac surgery, but its long-term impact is unclear.

**Methods:** Single-center retrospective analysis of individuals who underwent cardiac surgery from 2007-2022 measured the association of socioeconomic status and mortality. Mortality was determined by electronic health records and the National Death Index. Socioeconomic status was based on patients’ neighborhood Child Opportunity Index (COI) score. The lowest two COI quintile neighborhoods were designated “disadvantaged” while the highest two were designated “advantaged.” Multivariable mixed model analyses measured the strength of association between socioeconomic status and mortality, adjusting for surgical complexity (STAT category), demographic, and clinical factors.

**Results:** Of the 2,546 patients, half (49%, n=1,235) were from disadvantaged neighborhoods. One third (31%, n=787) were considered advantaged. Compared to patients from advantaged neighborhoods, patients from disadvantaged neighborhoods suffered greater overall mortality (14% vs 8%, p<0.001), more frequent complications (14% vs 10%, p<0.001), more genetic syndromes (23% vs 19%, p=0.021), and were smaller at the time of surgery (5.15 kg vs 5.60 kg, p=0.006). Multivariable analysis adjusting for STAT category, prematurity, weight at surgery, and presence of genetic syndromes found patients from disadvantaged neighborhoods had greater overall mortality (aOR:1.52; CI:1.10 – 2.12, p=0.013). Genetic syndromes and weight at surgery were associated with increased overall mortality (aOR:1.60; CI:1.17 – 2.20, p=0.004, aOR: 0.66; CI:0.53 – 0.81, p<0.001, respectively).

**Conclusions:** Patients from disadvantaged neighborhoods carry greater risk of mortality after pediatric cardiac surgery. The relationship between socioeconomic status and mortality may be mediated by weight and genetic syndromes and may accumulate over time.

## Introduction

Individuals from socioeconomically disadvantaged backgrounds experience disproportionate frequency of adverse healthcare outcomes. Such disparities persist in pediatric cardiac surgical care where disadvantaged patients experience longer lengths of stay, increased healthcare costs, and higher rates of reintervention after surgery^1,2^. Most seriously, previous literature has suggested that patients from lower socioeconomic backgrounds are at higher risk of operative mortality after pediatric cardiac surgery^3–6^.

Greater mortality in socioeconomically disadvantaged patients after cardiac surgery remains poorly understood. Furthermore, there is limited information on the impact of socioeconomic status (SES) on mortality beyond the immediate post-operative period. It is likely that current risk models in congenital heart surgery severely lack inclusion of patient factors such as the quality of the maternal fetal environment or sociodemographic factors^7^, and research suggests SES may be useful in models assessing longitudinal mortality risk in congenital heart disease surgery^8^. The objective of this study is to investigate the association between SES and risk of overall mortality after pediatric cardiac surgery. Secondary analysis examines the extent to which increased overall mortality is due to late mortality in disadvantaged populations.

## Methods

We performed a single-center retrospective analysis of individuals who underwent surgery to manage congenital cardiac disease over a 15-year period (January 1, 2007 to June 30, 2022). Only initial defect correcting surgeries were included for analysis, thus excluding cases such as isolated pacemaker implantations, ventricular assist device implantations, isolated patent ductus arteriosus closures or re-operations. In addition, individuals with unknown address or those living outside of the Washington, D.C. metropolitan area, as defined by the U.S. Office of Management and Budget, were excluded. The design of the study was approved by the ethics committee and a waiver of consent was obtained for the acquisition of data from the Institutional Review Board.

Clinical and demographic characteristics were collected from the electronic medical record, including cardiac diagnosis, gestational age at birth, genetic abnormalities (presence of either a known syndrome or chromosomal abnormality), non-cardiac abnormalities, presence of prenatal diagnosis, weight, height and age at surgery, surgical procedure, cross clamp and hypothermic arrest times, postoperative length of stay (PLOS), major surgical complications, and known mortality status, and Society of Thoracic Surgeons-European Association for Cardio-Thoracic Surgery (STAT) category. STAT category grades surgical risk (1 is the lowest, 5 the highest) and was used as a proxy for surgical complexity. Demographic characteristics collected included patient sex, ethnicity, race, insurance type and last known patient address.

### Childhood Opportunity Index Categorization

Childhood Opportunity Index (COI) is a composite metric of health, environment, education, social and economic factors that is used to reflect the socioeconomic background of a neighborhood^9^. COI ranks neighborhoods into five quintiles of “very low,” “low,” “moderate,” “high,” and “very high”. Each study patient was assigned a metropolitan-normed COI using the geocoded address to the nearest census tract, obtained from the publicly available COI 3.0 dataset. Study census tracts were categorized into disadvantaged COI (consisting of the very low and low quintiles) or advantaged COI (consisting of high and very high quintiles). The moderate quintile was ungrouped.

### Determination of Mortality Status

Those with known mortality were categorized as such. Any remaining individuals seen within the 18 months prior to data collection were considered alive. Any individual with no recent encounter had their mortality status validated by the National Death Index (NDI) from the Centers for Disease Control. Patient records were linked with data from the NDI, a reliable repository for tracking death records in the United States, using name, date of birth, and other demographic details, following strict data privacy and security protocols^10^. Matches were identified based on probabilistic linkage criteria, ensuring accuracy while addressing potential discrepancies (e.g., name variations or missing data)^11^. To further validate the linkage, we employed sensitivity checks, such as cross-referencing dates of death with other available clinical records and excluding ambiguous matches. Time of death was either categorized as operative mortality, occurring within 30 days or during initial hospitalization, or late mortality, defined as any occurring beyond 30 days or after hospital discharge (Table 1).

**Table 1:**
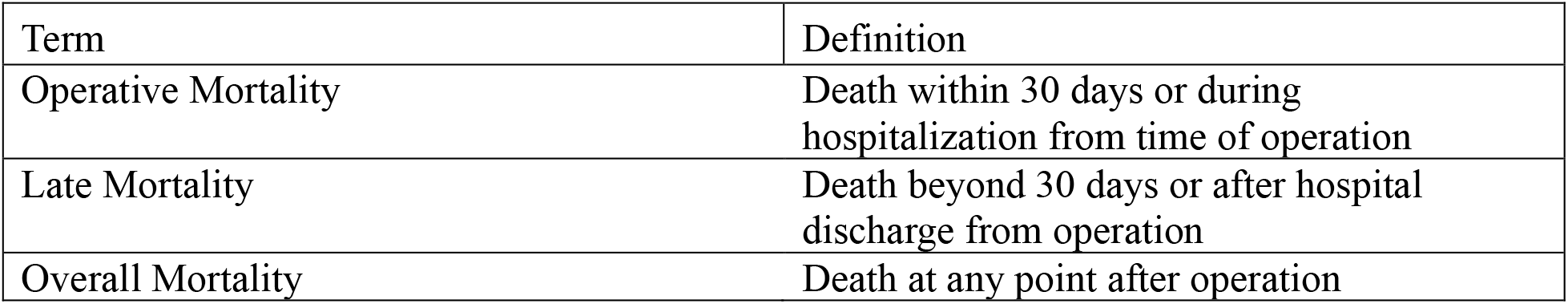
Mortality Definitions.

| Term | Definition |
| --- | --- |
| Operative Mortality | Death within 30 days or during hospitalization from time of operation |
| Late Mortality | Death beyond 30 days or after hospital discharge from operation |
| Overall Mortality | Death at any point after operation |

### Statistical Analyses

Patient demographics, surgical characteristics, and mortality outcomes were compared between COI neighborhoods. All categorical variables were assessed by chi-square or Fischer exact tests as appropriate while all the continuous variables were evaluated using t-tests or Kruskal Wallis tests as appropriate. To assess and compare the mortality outcomes, mixed model multivariable logistic regression analysis was performed. Patient demographic and clinical characteristics were adjusted as fixed effects for the models. Multivariable model selection was based on Lemeshow and Hosmer technique where all parameters, clinically and statistically significant, were selected and then forward-selection was used to identify the best models. Model selection was based on Akaike information criterion (AIC) or Bayesian information criterion (BIC) as appropriate. The accuracy of the model was assessed by C-statistic and area under the curve. For all the models, the assumptions and model fit were assessed, and necessary transformations were performed as needed. P-values <0.05 were considered statistically significant. Analyses were performed using SAS 9.4 software (SAS, Cary, NC, USA).

## Results

A total of 2,546 individuals who met inclusion criteria (breakdown of patient selection shown in Figure 1). The majority, 45% (n=1,081), were White while 36% (n=859) were Black and 19% (n=426) were in other race categories. Approximately half, 49% (n=1,235), of individuals resided in disadvantaged neighborhoods while 31% of individuals resided in advantaged neighborhoods (Table 2). More than half were male and median weight at time of surgery was 5.36 kg (IQR: 3.52 – 14.90). The study population was predominantly neonates and infants with 26% (n= 656) neonates, 37% (n=952) infants, 31% (n=785) children, and 6% (n=153) adults. Approximately half of the study population were classified as STAT Category 1, while 13% were categorized as STAT Category 4 or higher. A total of 16% (n=387) of individuals were premature (under 37 weeks gestational age) and one quarter (24%, n=609) had a genetic abnormality. Individuals from disadvantaged neighborhoods were smaller and younger at the time of surgery as compared to those from moderate and advantaged neighborhoods with the median height of 59cm (IQR: 51 – 91; p=0.007), median weight of 5.15 kg (IQR: 3.40 – 13.50; p=0.006), and youngest median age at surgery of 122 days (IQR: 21 – 1046; p=0.047). In general, individuals from disadvantaged neighborhoods experienced worse surgical conditions with longer cardiopulmonary bypass times (median: 102 min, IQR: 77 – 140; p<0.001), longer cross clamp times (51 min, IQR: 31 – 74; p<0.001), and experienced a higher proportion of urgent surgeries (29%, p=0.007). The amount of time spent in the hospital differed by socioeconomic status with individuals from disadvantaged neighborhoods having the longest postoperative lengths of stay at a median of 9 days (IQR: 5 – 22; p<0.001). Individuals from disadvantaged neighborhoods experienced higher rates of major complications (p<0.001), as well as higher rates of overall mortality (p<0.001) after cardiac surgery. In addition, the median time to death for individuals from disadvantaged neighborhoods who suffered from late mortality was shorter than those from advantaged neighborhoods (median: 485 days, IQR: 233 – 1441 vs. 523, IQR: 164 – 1346; p=0.001).

**Table 2:** Patient Demographics.

| Variable | Descriptor | Total | Disadvantaged<br>COI | Moderate<br>COI | Advantaged<br>COI | p-value |
| --- | --- | --- | --- | --- | --- | --- |
| Total Population (N=2,546) |  | 2546 (100%) | 1235 (48.5%) | 524<br>(20.6%) | 787 (30.9%) |  |
| Sex (n=2,546) |  |  |  |  |  | 0.780 |
|  | Female | 1152(45.2%) | 567(45.9%) | 236(45.0%) | 349(44.3%) |  |
|  | Male | 1394(54.8%) | 668(54.1%) | 288(55.0%) | 438(55.7%) |  |
| Race |  |  |  |  |  | <0.001 |
|  | White | 1080<br>(45.4%) | 381 (33.7%) | 221<br>(44.7%) | 478(63.2%) |  |
|  | Black | 858 (36.0%) | 565 (50.0%) | 180<br>(36.4%) | 113 (14.9%) |  |
|  | Other | 443 (18.6%) | 185 (16.4%) | 93 (18.8%) | 165 (21.8%) |  |
|  | Missing/Unknown | 165 | 104 | 30 | 31 |  |
| Insurance |  |  |  |  |  | <0.001 |
|  | Government | 791(48.5%) | 532(66.8%) | 136(41.5%) | 123(24.2%) |  |
|  | Commercial | 754(46.2%) | 229(28.8%) | 172(52.4%) | 353(69.5%) |  |
|  | Other | 37(2.3%) | 13(1.6%) | 13(4.0%) | 11(2.2%) |  |
|  | Multiple Insurances | 47(2.9%) | 21(2.6%) | 7(2.1%) | 19(3.7%) |  |
|  | No Insurance | 3(0.2%) | 1(0.1%) | 0 (0%) | 2(0.4%) |  |
|  | Missing/Unknown | 914 | 439 | 196 | 279 |  |
| Operative Type |  |  |  |  |  | 0.780 |
|  | Cardiopulmonary<br>Bypass | 2212(86.9%) | 1071(86.7%) | 460(87.8%) | 681(86.5%) |  |
|  | No Cardiopulmonary<br>Bypass | 334(13.1%) | 164(13.3%) | 64(12.2%) | 106(13.5%) |  |
| Operative<br>Status |  |  |  |  |  | 0.007 |
|  | Elective | 1843(73.0%) | 849(69.4%) | 391(75.0%) | 603(77.5%) |  |
|  | Emergent | 34(1.3%) | 15(1.2%) | 8(1.5%) | 11(1.4%) |  |
|  | Salvage | 3(0.1%) | 2(0.2%) | 0 (0%) | 1(0.1%) |  |
|  | Urgent | 643(25.5%) | 358(29.2%) | 122(23.4%) | 163(21.0%) |  |
|  | Missing/Unknown | 23 | 11 | 3 | 9 |  |
| Overall Mortality |  |  |  |  |  | <0.001 |
|  | No | 2274(89.3%) | 1067(86.4%) | 486(92.7%) | 721(91.6%) |  |
|  | Yes | 272(10.7%) | 168(13.6%) | 38(7.3%) | 66(8.4%) |  |
| Major Complication |  |  |  |  |  | <0.001 |
|  | No | 2258(88.7%) | 1067(86.4%) | 485(92.6%) | 706(89.7%) |  |
|  | Yes | 288(11.3%) | 168(13.6%) | 39(7.4%) | 81(10.3%) |  |
| Age Category |  |  |  |  |  | 0.050 |
|  | Adult (>18 yrs) | 153(6.0%) | 70(5.7%) | 28(5.3%) | 55(7.0%) |  |
|  | Child (1–18 yrs) | 785(30.8%) | 349(28.3%) | 179(34.2%) | 257(32.7%) |  |
|  | Infant (3mo – 1 yr) | 952(37.4%) | 476(38.5%) | 199(38.0%) | 277(35.2%) |  |
|  | Neonate (0-3mo) | 656(25.8%) | 340(27.5%) | 118(22.5%) | 198(25.2%) |  |
| Reoperation<br>Type |  |  |  |  |  | 0.740 |
|  | None | 2516(99.3%) | 1218(99.3%) | 519(99.4%) | 779(99.2%) |  |
|  | Yes - Planned | 11(0.4%) | 4(0.3%) | 2(0.4%) | 5(0.6%) |  |
|  | Yes - Unplanned | 6(0.2%) | 4(0.3%) | 1(0.2%) | 1(0.1%) |  |
|  | Missing/Unknown | 13 | 9 | 2 | 2 |  |
| STAT Category |  |  |  |  |  | 0.017 |
|  | 1 | 1362(54.1%) | 617(50.5%) | 299(57.6%) | 446(57.3%) |  |
|  | 2 | 551(21.9%) | 270(22.1%) | 108(20.8%) | 173(22.2%) |  |
|  | 3 | 286(11.4%) | 162(13.3%) | 51(9.8%) | 73(9.4%) |  |
|  | 4 | 188(7.5%) | 98(8.0%) | 40(7.7%) | 50(6.4%) |  |
|  | 5 | 132(5.2%) | 74(6.1%) | 21(4.0%) | 37(4.7%) |  |
|  | Missing/Unknown | 27 | 14 | 5 | 8 |  |
| Antenatal Diagnosis |  |  |  |  |  | 0.033 |
|  | No | 1517 (64.2%) | 759 (66.3%) | 315 (64.7%) | 443 (60.4%) |  |
|  | Yes | 847 (35.8%) | 385 (33.7%) | 172 (35.3%) | 290 (39.6%) |  |
|  | Missing/Unknown | 182 | 91 | 37 | 54 |  |
| Premature Birth |  |  |  |  |  | 0.300 |
|  | No | 2011(83.9%) | 975(83.2%) | 400(82.8%) | 636(85.6%) |  |
|  | Yes | 387(16.1%) | 197(16.8%) | 83(17.2%) | 107(14.4%) |  |
| Non-Cardiac Abnormalities |  |  |  |  |  | 0.590 |
|  | No | 2270(89.2%) | 1095(88.7%) | 466(88.9%) | 709(90.1%) |  |
|  | Yes | 276(10.8%) | 140(11.3%) | 58(11.1%) | 78(9.9%) |  |
| Syndromes |  |  |  |  |  | 0.012 |
|  | No | 2015(79.1%) | 947(76.7%) | 428(81.7%) | 640(81.3%) |  |
|  | Yes | 531(20.9%) | 288(23.3%) | 96(18.3%) | 147(18.7%) |  |
| Chromosomal Abnormalities |  |  |  |  |  | 0.030 |
|  | No | 2045(80.3%) | 966(78.2%) | 435(83.0%) | 644(81.8%) |  |
|  | Yes | 501(19.7%) | 269(21.8%) | 89(17.0%) | 143(18.2%) |  |
| Genetic Abnormalities |  |  |  |  |  | 0.021 |
|  | No | 1937(76.1%) | 909(73.7%) | 411(78.4%) | 617(78.4%) |  |
|  | Yes | 609(23.9%) | 324(26.3%) | 113(21.6%) | 170(21.6%) |  |
| Weight at Surgery (kg) |  | 5.36 (3.52 – 14.90) | 5.15 (3.40 – 13.50) | 5.63 (3.69 – 16.15) | 5.60 (3.60 – 16.50) | 0.006 |
| Height at Surgery (cm) |  | 60 (51 – 96) | 59 (51 – 91) | 60 (52 – 100.5) | 61 (52 – 101.5) | 0.007 |
| Age at Operation (days) |  | 136 (25 –<br>1322) | 122 (21 –<br>1046) | 156 (41 –<br>15673) | 145 (29 –<br>1564) | 0.047 |
| Hospital Length of Stay (days) |  | 9 (5 – 26) | 12 (5 – 32) | 8 (4 – 23) | 7 (4 – 20) | <0.001 |
| Postoperative Length of Stay (days) |  | 7 (4 – 19) | 9 (5 – 22) | 6 (4 – 16) | 6 (4 – 15) | <0.001 |
| Cardiopulmonary Bypass Time (min) |  | 96 (72 –<br>133) | 102 (77 – 140) | 90 (70 –<br>125) | 87 (68 –<br>123) | <0.001 |
| Cross Clamp Time (min) |  | 48 (30 – 70) | 51 (31 – 74) | 44 (31 –<br>65) | 45 (29 – 66) | <0.001 |

COI: Child Opportunity Index
STAT: Society of Thoracic Surgeons-European Association for Cardio-Thoracic Surgery

**Figure 1:**
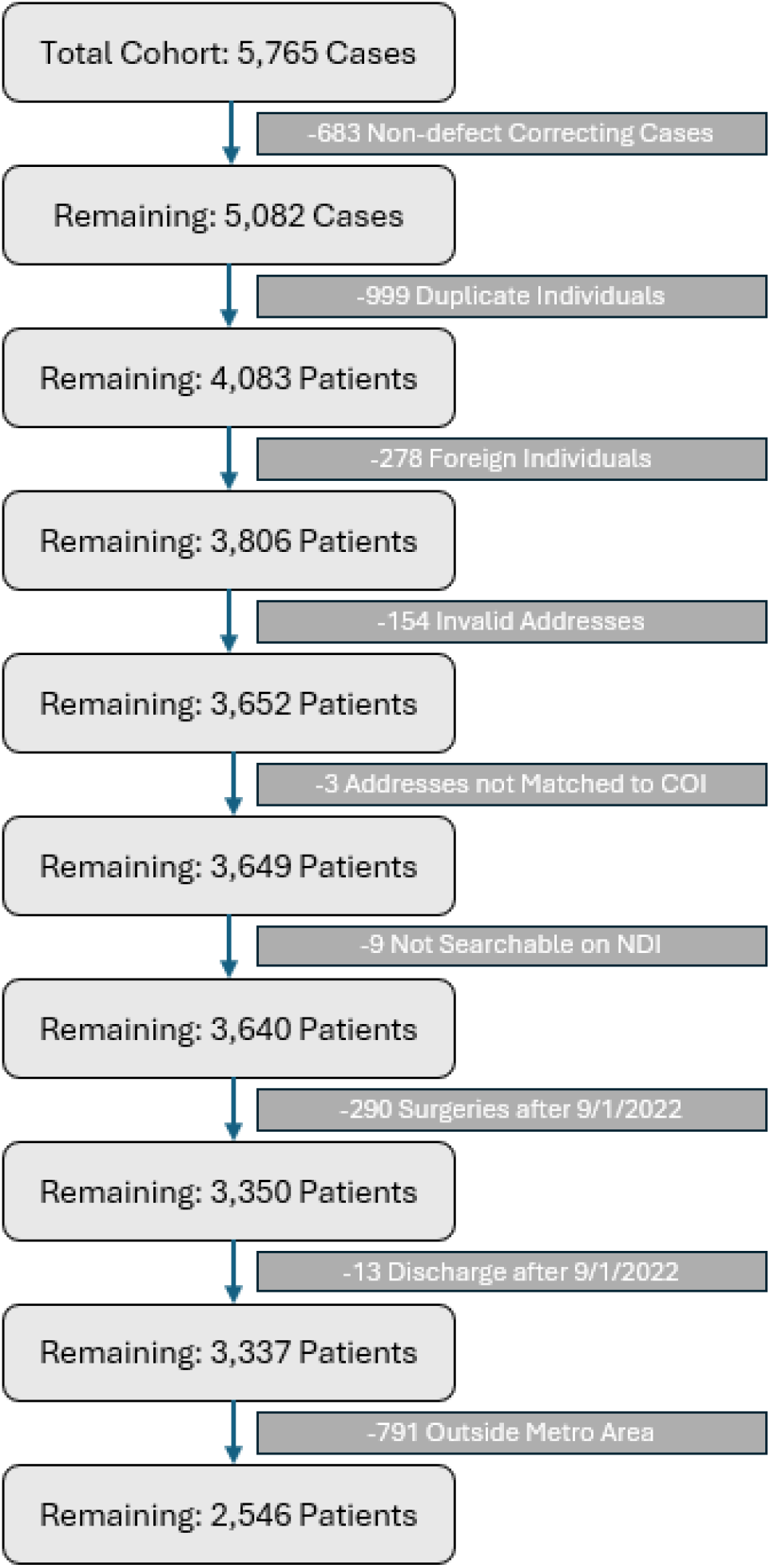
Application of inclusion and exclusion criteria to the initial potential cohort of 5,765 cases. Only the first defect-correcting case was included and those who could not be socioeconomically grouped by COI or have mortality status determined were excluded. COI: Child Opportunity Index NDI: National Death Index

Bivariable analysis assessed demographic and clinical factors associated with overall mortality after cardiac surgery (Table 3). Individuals from disadvantaged neighborhoods had 1.7 times higher odds (95% CI: 1.27 – 2.32) of mortality as compared to those from advantaged neighborhoods (p<0.001). Notably, this relationship did not exist for individuals from moderate socioeconomic neighborhoods compared to those from advantaged neighborhoods (p=0.205). Baseline clinical factors and surgical details also conveyed risk for mortality after cardiac surgery. Neonates, designated as younger than 3 months, had nearly eight times higher odds of mortality in comparison with older children who underwent cardiac surgery (aOR: 7.81, 95% CI: 5.13 – 11.88, p<0.001). Individuals born prematurely had two times higher odds of mortality in comparison with those that were full-term (aOR: 2.09, 95% CI: 1.56 – 2.81, p<0.001). Individuals with chromosomal abnormality, genetic issues, and lower weight also had higher likelihood of mortality in comparison with those that did not have these conditions. Surgical factors, including longer bypass and cross-clamp times, were also associated with increased mortality risk. Surgical STAT categories 2 – 5 had higher odds of mortality in comparison with the STAT category 1, underscoring the risk associated with increasing complexity of operation.

**Table 3:** Bivariable Analysis of Overall Mortality.

| Variable | Odds Ratio (95% Confidence Interval) | p-value |
| --- | --- | --- |
| Sex (Female vs Male) | 1.24 (0.96 – 1.59) | 0.096 |
| Operative Type<br>(Cardiopulmonary bypass vs<br>No Cardiopulmonary bypass) | 0.41 (0.30 – 0.55) | <.001 |
| Age (Adult vs Child) | 1.89 (0.90 – 3.98) | 0.257 |
| Age (Infant vs Child) | 2.72 (0.76 – 4.21) | 0.550 |
| Age (Neonate vs Child) | 7.81 (5.13 – 11.88) | <0.001 |
| STAT Category 2 vs 1 | 2.85 (1.90 – 4.28) | <.001 |
| STAT Category 3 vs 1 | 6.81 (4.53 – 10.26) | 0.040 |
| STAT Category 4 vs 1 | 9.65 (6.25 – 14.91) | <.001 |
| STAT Category 5 vs 1 | 22.13 (14.14 – 34.64) | <.001 |
| Premature Birth (yes vs no) | 2.09 (1.56 – 2.81) | <.001 |
| Non-cardiac Abnormality<br>(yes vs no) | 1.70 (1.20 – 2.41) | 0.003 |
| Syndromes (yes vs no) | 1.72 (1.30 – 2.27) | 0.001 |
| Chromosomal abnormality<br>(yes vs no) | 1.52 (1.13 – 2.03) | 0.005 |
| Genetic Abnormalities (yes vs<br>no) | 1.71 (1.31 – 2.24) | <.001 |
| COI (Disadvantaged vs<br>Advantaged) | 1.72 (1.27 – 2.32) | <.001 |
| COI (Disadvantaged vs<br>Moderate) | 2.01 (1.39 – 2.91) | <.001 |
| COI (Moderate vs<br>Advantaged) | 0.85 (0.56 – 1.29) | 0.205 |
| Weight at Surgery | 0.38 (0.31 – 0.47) | <.001 |
| Cardiopulmonary Bypass Time | 5.33 (3.77 – 7.53) | <.001 |
| Cross Clamp Time | 2.07 (1.51 – 2.85) | <0.01 |
| Hospital Length of Stay | 1.02 (1.01 – 1.02) | <0.01 |
| Postop Length of Stay | 1.02 (1.02 – 1.02) | <.001 |

COI: Child Opportunity Index
STAT: Society of Thoracic Surgeons-European Association for Cardio-Thoracic Surgery

Multivariable analyses revealed associations for both overall and late mortality, highlighting the long-term interplay of socioeconomic, clinical, and surgical factors (Tables 4 and 5). Individuals from disadvantaged communities exhibited 1.6 times higher odds of late mortality compared to those from advantaged communities (aOR: 1.58, 95% CI: 1.03 – 2.42, p = 0.038). Similar trends held true for overall mortality, with those from disadvantaged areas experiencing 1.5 times higher odds of mortality at any point after surgery (aOR: 1.52, 95% CI: 1.10 – 2.12, p = 0.013). Of note, higher STAT category, prematurity, low weight at surgery, and presence of genetic syndromes or abnormalities were significant predictors of overall mortality. The risk of mortality increased over time for individuals for disadvantaged neighborhoods; the 15-year freedom from mortality for individuals from disadvantaged neighborhoods was lower than those from advantaged neighborhoods (81% vs. 90%; p<0.001) (Figure 2).

**Table 4:** Multivariable Analysis (Overall Mortality)

| Variable | Adjusted Odds Ratio (95% Confidence Interval) | p-value |
| --- | --- | --- |
| Disadvantaged vs. Advantaged COI | 1.52 (1.10 – 2.12) | 0.013 |
| Moderate vs. Advantaged COI | 0.90 (0.57 – 1.40) | 0.608 |
| STAT Category (2 vs 1) | 2.40 (1.58 – 3.65) | <0.001 |
| STAT Category (3 vs 1) | 5.47 (3.56 – 8.40) | 0.065 |
| STAT Category (4 vs 1) | 6.59 (4.14 – 10.49) | 0.004 |
| STAT Category (5 vs 1) | 15.78 (9.70 – 25.66) | <0.001 |
| Prematurity (yes vs no) | 1.77 (1.26 – 2.49) | 0.001 |
| Syndrome (yes vs no) | 1.60 (1.17 – 2.20) | 0.004 |
| Weight at Surgery | 0.66 (0.53 – 0.81) | <.001 |

COI: Child Opportunity Index
STAT: Society of Thoracic Surgeons-European Association for Cardio-Thoracic Surgery

**Table 5:** Multivariable Analysis (Late Mortality)

| Variable | Adjusted Odds Ratio (95% Confidence Interval) | p-value |
| --- | --- | --- |
| Disadvantaged vs. Advantaged COI | 1.58 (1.03 – 2.42) | 0.038 |
| Moderate vs. Advantaged COI | 0.82 (0.45 – 1.51) | 0.524 |
| STAT Category (2 vs 1) | 2.27 (1.37 – 3.74) | 0.001 |
| STAT Category (3 vs 1) | 4.23 (2.52 – 7.12) | <0.001 |
| STAT Category (4 vs 1) | 3.46 (1.86 – 6.45) | <0.001 |
| STAT Category (5 vs 1) | 8.05 (4.51 – 14.36) | <0.001 |
| Syndrome (yes vs no) | 1.72 (1.16 – 2.55) | 0.008 |

COI: Child Opportunity Index
STAT: Society of Thoracic Surgeons-European Association for Cardio-Thoracic Surgery

**Figure 2:**
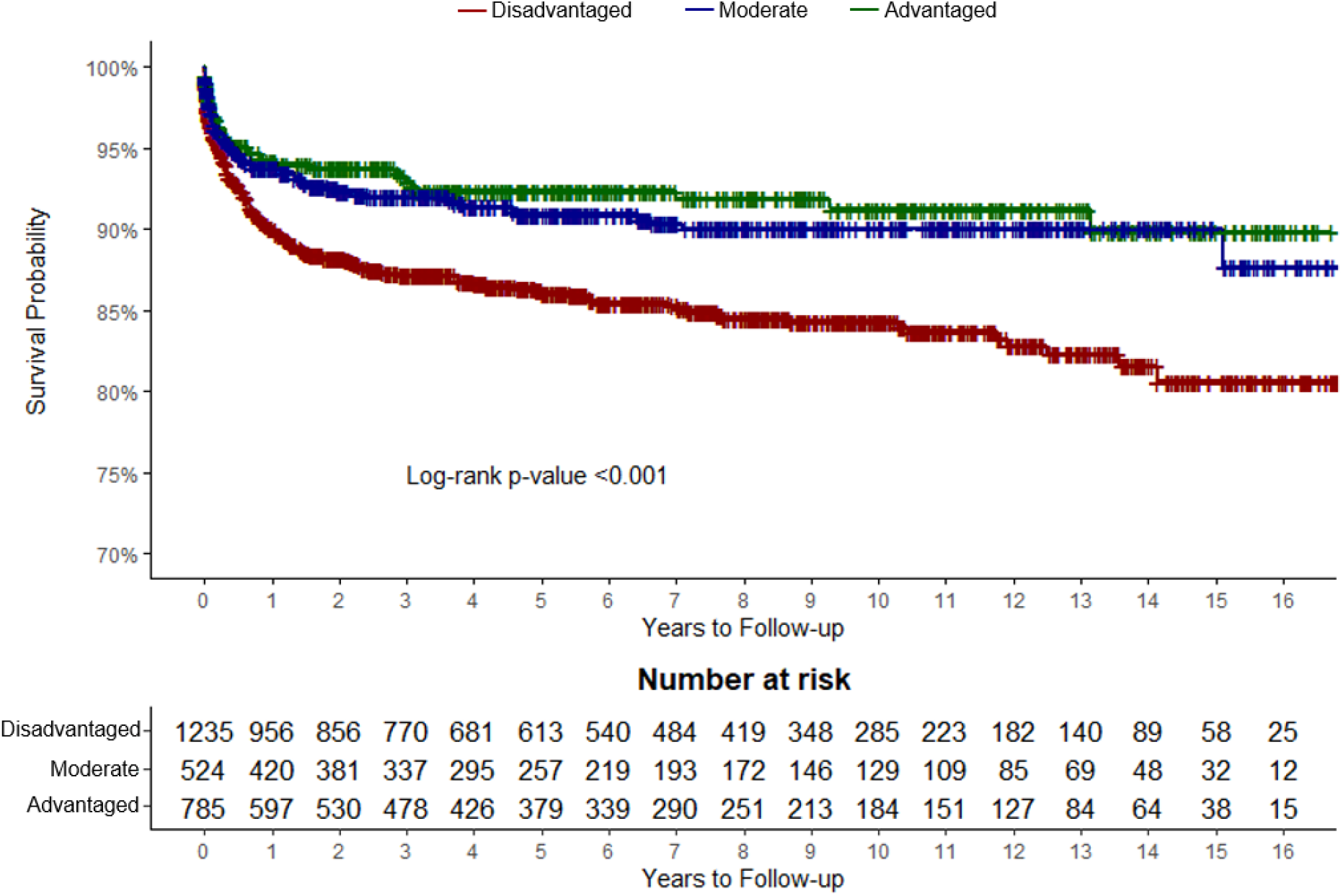
Overall Mortality by Neighborhood Childhood Opportunity Index Designation Kaplan-Meier curve analysis comparing the estimated risk of mortality over time between different socioeconomic groups. After a period of 15 years of follow up, the estimated survival odds of individuals from disadvantaged communities were significantly lower than the survival rates of those from moderate and advantaged communities. COI: Child Opportunity Index NDI: National Death Index

## Discussion

We previously described a 50 percent increase in the risk of operative mortality for patients from disadvantaged neighborhoods^3^, but with only borderline statistical significance, which was not present when adjusted for potential confounders. The current study finds the impact of neighborhood disadvantage is considerably more pronounced in late and overall mortality and holds true across multivariable models. In addition, it suggests that late mortality, as opposed to operative mortality, drives the overall mortality association with disadvantaged neighborhood residence.

Results from operative mortality, late mortality, and overall mortality analyses suggest a number of clinical and surgical factors may drive the association between neighborhood disadvantage and mortality after cardiac surgery. In particular, small size at surgery and the presence of a genetic abnormality are significantly increased for those from disadvantaged neighborhoods and are also associated with increased mortality across all models. While these associations may not be new – genetic abnormalities and smaller size carry a demonstrable increased risk of mortality after cardiac surgery^12–14^ – the link with socioeconomic disadvantage compounds the risk for already vulnerable individuals.

The primary limitation of this study is socioeconomic status extrapolated from the individual’s neighborhood rather than using individual socioeconomic metrics. Additionally, cause of death was not available for analysis. Future studies will investigate the cause of death for individuals after cardiac surgery to identify potential areas for intervention.

Alarmingly, the impact of socioeconomic background, as captured by neighborhood advantage, on mortality after congenital cardiac surgery appears to accumulate over time. Prior research has shown lower socioeconomic status to be associated with increased mortality risk later in life^15–18^, though to our knowledge this hasn’t been demonstrated in the population of those undergoing pediatric heart surgery. It has been suggested that health behaviors alone do not account for the increase in mortality observed^19,20^. Socioeconomic disparities in late mortality, potentially driven by clinical and surgical factors, requires further investigation and intervention.

## Data Availability

Due to protecting patients’ privacy and the sensitive nature of the research, data is not publicly available.

## Non-standard Abbreviations and Acronyms

AIC: Akaike Information Criterion
aOR: Adjusted Odds Ratio
BIC: Bayesian information criterion
COI: Child Opportunity Index
NDI: National Death Index
PLOS: Post Operative Length of Stay
SES: Socioeconomic Status
STAT: Society of Thoracic Surgeons-European Association for Cardio-Thoracic Surgery

## Acknowledgements

With thanks to In Hye Park for the instrumental support of this investigation.

